# Systemic adverse effects induced by the BNT162b2 vaccine are associated with higher antibody titers from 3 to 6 months after vaccination

**DOI:** 10.1101/2022.01.23.22269706

**Authors:** Ryousuke Koike, Michiru Sawahata, Yosikazu Nakamura, Yushi Nomura, Otohiro Katsube, Koichi Hagiwara, Seiji Niho, Norihiro Masuda, Takaaki Tanaka, Kumiya Sugiyama

## Abstract

**Objective:** We aimed to determine the relationship between vaccine-related adverse effects and antibody (Ab) titers from 3 to 6 months after the second dose of the BNT162b2 coronavirus disease 2019 (COVID-19) mRNA vaccine (Pfizer/BioNTech) in Japan.

**Methods:** We enrolled 378 healthcare workers (255 women, 123 men) whose Ab titers 3 and 6 months after the second dose were analyzed in our previous study and whose characteristics and adverse effects were collected previously using a structured self-report questionnaire.

**Results:** Median age was 44 years. While injection-site symptoms occurred with almost equal frequency between the first and second doses, systemic adverse effects, such as general fatigue and fever, were significantly more frequent after the second dose than after the first. Multivariate analysis showed that fever was significantly correlated with female sex for the second dose (odds ratio [OR], 2.139; 95% confidence interval [95%CI], 1.185–3.859), older age for the first dose (OR, 0.962; 95%CI, 0.931–0.994) and second dose (OR, 0.957; 95%CI, 0.936–0.979), and dyslipidemia for the first dose (OR, 8.750; 95%CI, 1.814–42.20). Age-adjusted Ab titers at 3 months after vaccination were 23.7% and 23.4% higher in patients with fever than in those without fever after first and second dose, respectively. In addition, age-adjusted Ab titers at 3 and 6 months after the second dose were respectively 21.7% and 19.3% higher in the group with anti-inflammatory agent use than in the group without anti-inflammatory agent use.

**Conclusion:** Participants with systemic adverse effects tend to have higher Ab titers from 3 to 6 months after the second dose of the BNT162b2 vaccine. Our results may encourage vaccination, even among people with vaccine hesitancy related to relatively common systemic adverse effects.

## INTRODUCTION

The BNT162b2 vaccine (Pfizer/BioNTech) was selected as the first coronavirus disease 2019 (COVID-19) mRNA vaccine to be administered to healthcare professionals in Japan, beginning in February 2021. The vaccine helps to prevent not only the onset [1], but also the progression of COVID-19. However, some people hesitate to get vaccinated due to fear of relatively common adverse effects that occur shortly after vaccination with the BNT162b2 vaccine [2-4].

If the maximum vaccine efficacy, including antibody (Ab) responses, is guaranteed, most people will accept a certain severity of vaccination-related adverse effects [2]. Although several reports concluded that there is no or only a low correlation between adverse effects and the Ab titers detected immediately after vaccination [5-8], a correlation is expected between adverse effects and the Ab titers detected several months after vaccination [9]. To examine the efficacy of vaccination, it is more important to analyze the moderate-term levels of Ab titers after vaccination rather than the peak Ab titer obtained shortly after vaccination. Indeed, several reports [6, 10] suggested that some systemic adverse effects may be induced as a hypersensitive immune reaction to the accumulation of severe acute respiratory syndrome coronavirus 2 (SARS-CoV-2) spike protein itself immediately after the injection of the first mRNA vaccine dose rather than chemical additives in vaccines. Analysis of the response to the first vaccine dose in persons with previous COVID-19 infection revealed that Ab titers were 10 to 45 times higher in individuals with preexisting immunity than in those without preexisting immunity at the same time points after the first vaccine dose [10]. Interestingly, systemic adverse effects, including fatigue, headache, chills, muscle pain, fever, and joint pain, were more common among participants with preexisting immunity while local adverse effects occurred with similar frequency among participants with and without preexisting immunity [10].

In our previous studies [11, 12], we showed that the most important factors associated with lower Ab titers at 3 and 6 months were age and smoking, probably reflecting their effect on peak Ab titers, and that the only factor significantly associated with the attenuation in Ab titer from 3 to 6 months was sex, which reduced the sex difference seen during the first 3 months. Against this background, we analyzed the relationship between adverse effects and Ab titers against the SARS-CoV-2 spike antigen from 3 to 6 months after vaccination with the BNT162b2 vaccine in Japan. This study is expected to contribute to maximizing the efficacy of vaccination while minimizing adverse effects as a step toward optimizing and individualizing the vaccination regimen.

## METHODS

### Population and study design

In our previous single-center prospective observational study [11, 12], we enrolled 378 healthcare workers (255 women, 123 men) at the National Hospital Organization Utsunomiya National Hospital in Tochigi Prefecture, Japan and analyzed their Ab titers at 3 and 6 months after the second of two BNT162b2 vaccine inoculations administered 3 weeks apart in February and March 2021. The participants’ characteristics and adverse effects were collected using a structured self-report questionnaire and, in this study, we analyzed the relationships of adverse effects with Ab titers at 3 and 6 months.

In our previous study [11], we used the participants’ blood samples to measure total Ab titers against the SARS-CoV-2 spike antigen using a commercially available electrochemiluminescence immunoassay (ECLIA) (Elecsys® Anti-SARS-CoV-2 RUO; Roche Diagnostics) [13]. In addition to their clinical and demographic characteristics, adverse effects occurring shortly after the first and second doses were recorded in that study using a structured self-report questionnaire [11]. In total, 365 healthcare workers (250 women, 115 men) remained after the exclusion of 10 participants whose blood samples were not obtained at 6 months and 3 participants whose blood sampling confirmed the presence of Abs against the nucleocapsid proteins for SARS-CoV-2 [12]; the blood samples of these 365 participants were used to measure total Ab titers against the SARS-CoV-2 spike antigen 6 months after the second dose. The relationships between Ab titers against the SARS-CoV-2 spike antigen and clinical and lifestyle characteristics were analyzed. In age-adjusted analysis, individual Ab titers were recalculated by subtracting the median Ab titer of the corresponding age group from an individual’s Ab titer. For example, an age-adjusted Ab titer for an individual in his/her 20s was calculated as follows: individual Ab titer – median Ab titer for participants in their 20s.

The Ethics Committee of National Hospital Organization Utsunomiya National Hospital (No. 03-01; April 19, 2021) approved this study, and written informed consent was obtained from all participants before their enrollment.

### Structured self-report questionnaire to obtain individual variables

Adverse effects in response to the first and second doses of the BNT162b2 vaccine, as well as clinical history and demographic characteristics, were collected by means of a structured self-report questionnaire. The maximum body temperature induced by the BNT162b2 vaccine was included in the questionnaire. In addition, we asked participants about the use of an anti-inflammatory agent for adverse effects induced by the BNT162b2 vaccine. However, preventive use of medication was excluded.

### Data analysis

Nonparametric continuous data are expressed as the median with the interquartile range (IQR). Categorical data are presented as absolute numbers (*n*) and relative frequencies (%). We used Microsoft^®^ Excel^®^ 2016 MSO (Microsoft Corp., Redmond, WA). To calculate Spearman’s rank correlation coefficient and perform the Mann–Whitney *U* test, chi-square test, and multivariate logistic regression analysis, we used the Statistical Package for Social Sciences (SPSS) version 28 (IBM Japan, Ltd., Tokyo, Japan). Univariate and multivariate logistic regression models were applied to analyze the relationship between the SARS-CoV-2 Ab or fever as the dependent variable and adverse effects or clinical parameters as the independent variables.

## RESULTS

### Incidence of adverse effects in response to the BNT162b2 vaccine

The participants’ baseline characteristics were reported in our previous publications [11, 12]. Overall, the vaccine had no adverse effects resulting in hospitalization. The prevalence of adverse effects and anti-inflammatory agent use in response to the BNT162b2 vaccine is shown in Table 1. The prevalence of each was higher after the second vaccine dose than after the first dose (chi-square test, *P*<0.01). Local adverse effects occurred with almost equal frequency for the first and second doses. The rates of concordance, in which the same reactions were seen for both the first and second doses, were higher for systemic reactions than for local reactions.

**Table 1.** Prevalence of adverse effects induced by the BNT162b2 vaccine (n=378)

|  | After first dose | After second dose | Rate of concordance<br>between 1<sup>st</sup> and 2<sup>nd</sup> |
| --- | --- | --- | --- |
| **Systemic reactions** |  |  |  |
| Fever $\geq 37.0^{\circ}\text{C}$ | 17.0% (56/274/48) | 56.8% (191/145/42) | 83.9% (47/56) |
| General fatigue | 35.0% (132/245/1) | 73.3% (277/101/0) | 93.9% (124/132) |
| Headache | 20.7% (78/299/1) | 46.0% (173/203/2) | 79.5% (62/78) |
| Muscle pain | 18.6% (70/307/1) | 35.7% (135/243/0) | 80.0% (56/70) |
| Joint pain | 7.7% (29/348/1) | 33.2% (125/252/1) | 86.2% (25/29) |
| Nausea | 2.4% (9/368/1) | 7.0% (26/347/5) | 66.7% (6/9) |
| Diarrhea | 1.1% (4/373/1) | 3.7% (14/362/2) | 50.0% (2/4) |
| **Local reactions** |  |  |  |
| Pain | 89.6% (337/39/2) | 83.3% (310/62/6) | 87.2% (294/337) |
| Swelling | 31.6% (119/257/2) | 31.1% (117/259/2) | 74.8% (89/119) |
| Induration | 23.4% (88/288/2) | 21.0% (79/297/2) | 72.7% (64/88) |
| Itching | 16.5% (62/314/2) | 17.2% (65/313/0) | 72.6% (45/62) |
| Medication use | 21.1% (76/284/18) | 57.1% (210/158/10) | 76.3% (58/76) |

Data show the percentage of positive reactions (number of yes/no/unknown)

### Relationship between participants’ characteristics and vaccine-related systemic adverse effects

Fever is one of the most severe systemic adverse effects, and we used multivariate logistic regression analysis to investigate whether participants’ characteristics were risk factors for the occurrence of fever (**Table 2**). Fever was defined as a maximum body temperature of 37.0LJ or higher. The multivariate analysis found that female sex (odds ratio [OR], 2.139; 95% confidence interval [95%CI], 1.185–3.859) was correlated with the occurrence of fever after the second dose. Younger age was also correlated with the occurrence of fever after the first dose (OR, 0.962; 95%CI, 0.931–0.994) and second dose (OR, 0.957; 95%CI, 0.936–0.979). A scatter plot of the distribution of body temperature by age showed that younger participants had a significantly higher body temperature shortly after both the first and second doses (correlation coefficient ρ=−0.370 and −0.184, respectively) (**Figure 1**). Dyslipidemia was also correlated with fever after the first dose (OR, 8.750; 95%CI, 1.815–42.25), but no other clinical or lifestyle characteristics were correlated with fever.

**Table 2.**
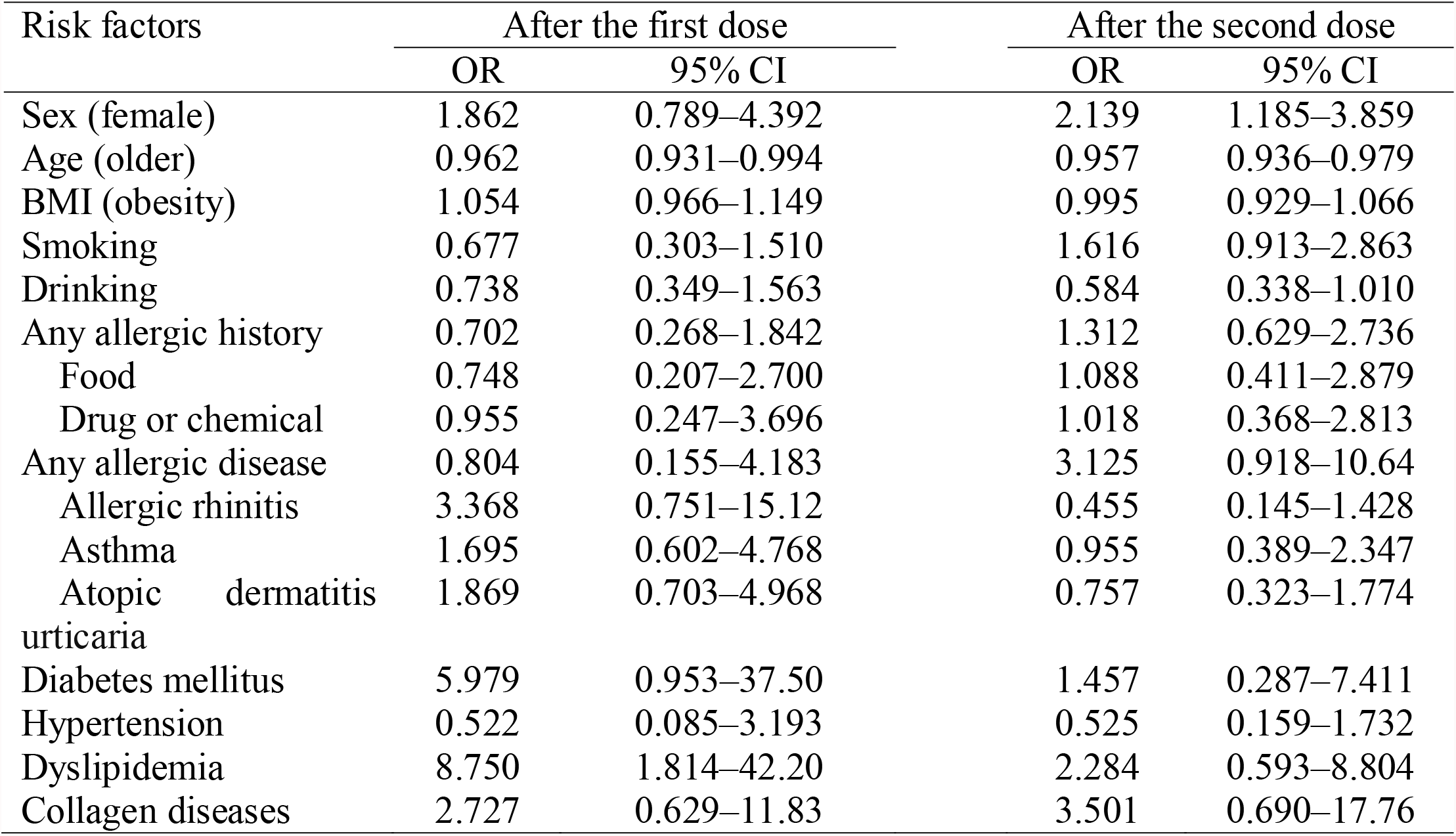
Multivariate logistic regression analysis of risk factors associated with fever (≥37.0°C) (n=378)

**Figure 1.**
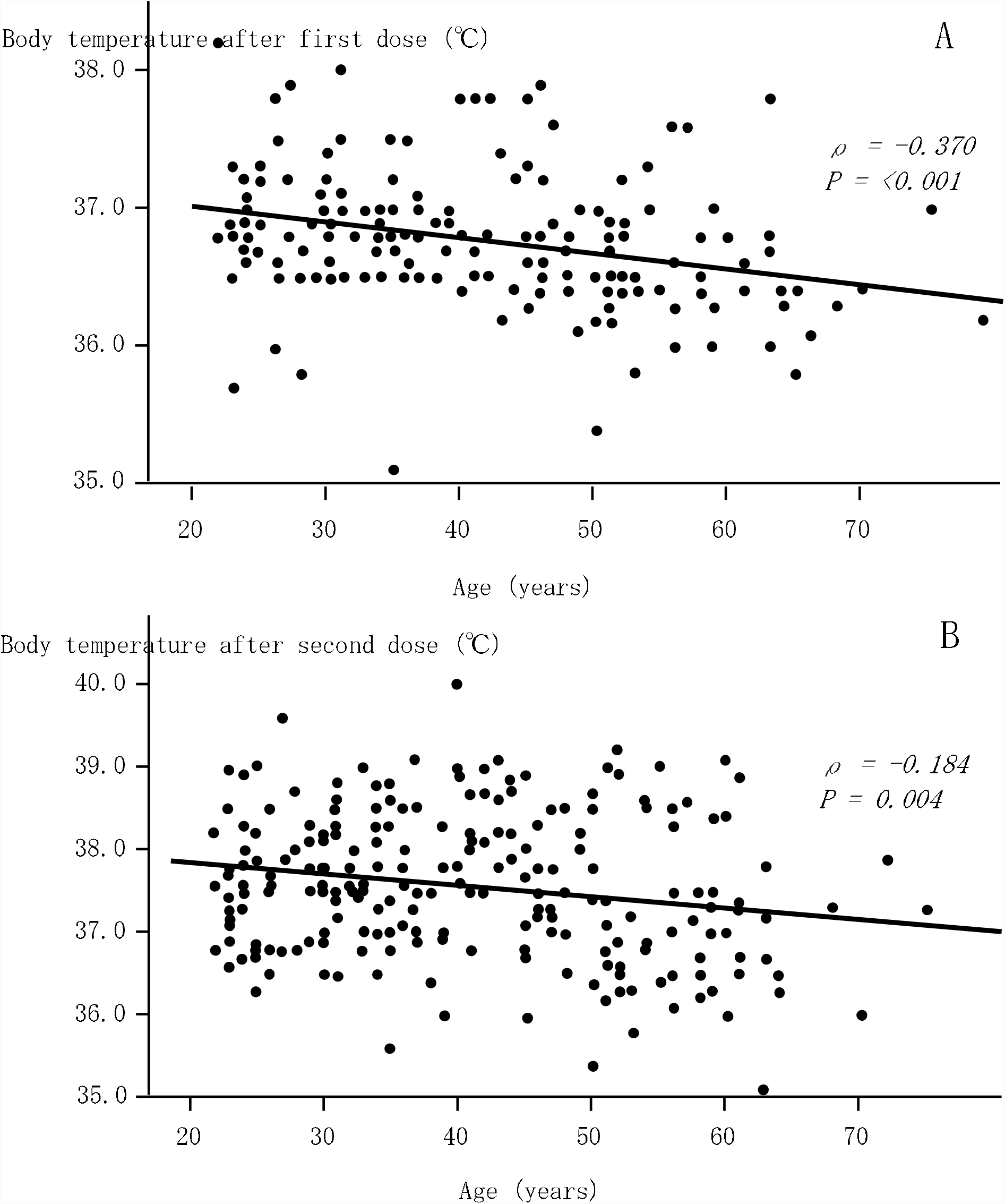
Relationships between age and body temperature after the first (A) and second (B) vaccine doses. Significant negative correlations were observed after both the first and second vaccine doses.

### Association between Ab titers and vaccine-related adverse effects

We analyzed the relationship between Ab titers at 3 and 6 months after vaccination and adverse effects after the first and second vaccine doses (**Tables 3 and 4**). Age-adjusted Ab titers were used because the titers significantly differed between younger and older participants at 3 and 6 months after vaccination. Fever after both the first and second doses was associated with a significantly higher Ab titer at 3 months after vaccination. A scatter plot of the distribution of body temperature by Ab titer also showed that participants with a high body temperature as an adverse effect after the second dose had a significantly higher Ab titer at 3 months after vaccination but not at 6 months (correlation coefficients ρ=0.166 and −0.102, respectively) (**Figure 2**). General fatigue and joint pain after the first and/or second vaccine doses were associated with significantly higher Ab titers at both 3 and 6 months after vaccination. Nausea after the second vaccine dose was associated with a significantly higher Ab titer at 6 months after vaccination. In addition, use of an anti-inflammatory agent after the second vaccine dose was significantly associated with high Ab titers at both 3 and 6 months after vaccination. However, local reactions were not associated with Ab titers.

**Table 3.** Relationship between adverse effects and age-adjusted antibody titers at 3 months after vaccination (n=378)

|  | After the first dose<br>Median (IQR) U/mL | *P*-value | After the second dose<br>Median (IQR) U/mL | *P*-value |
| --- | --- | --- | --- | --- |
| Systemic reactions |  |  |  |  |
| Fever ($\geq 37.0^{\circ}\text{C}$) | 153 (–242 to 529) / –28 (–306 to 293) | 0.042 | 110 (–259 to 511) / –69 (–335 to 263) | 0.002 |
| General fatigue * | 22 (–337 to 369) / 0 (–279 to 334) | 0.841 | 60 (–287 to 430) / –109 (–327 to 130) | 0.010 |
| Headache * | –30 (–416 to 334) / 0 (–285 to 355) | 0.289 | 70 (–309 to 481) / –47 (–293 to 297) | 0.120 |
| Muscle pain * | 32 (–329 to 395) / 0 (–293 to 341) | 0.770 | 90 (–246 to 340) / –39 (–336 to 352) | 0.058 |
| Joint pain * | 189 (–133 to 751) / –13 (–308 to 321) | 0.014 | 147 (–140 to 510) / –72 (–370 to 275) | <0.001 |
| Nausea * | 145 (–516 to 369) / 0 (–299 to 340) | 0.926 | –249 (–441 to 171) / 9 (–285 to 353) | 0.088 |
| Diarrhea * | 418(120 to 691) / 0 (–299 to 340) | 0.295 | –91 (–413 to 376) / 1 (–296 to 341) | 0.430 |
| Local reactions |  |  |  |  |
| Pain * | 2 (–308 to 369) / –88 (–256 to 237) | 0.498 | –1 (–317 to 341) / 26 (–226 to 335) | 0.528 |
| Swelling * | 0 (–307 to 395) / –3 (–299 to 317) | 0.822 | 0 (–312 to 411) / –11 (–293 to 322) | 0.780 |
| Induration * | 38 (–293 to 510) / –1 (–299 to 318) | 0.289 | –18 (–311 to 316) / 0 (–299 to 339) | 0.902 |
| Itching * | 81 (–174 to 451) / –17 (–309 to 339) | 0.265 | 9 (–273 to 269) / 0 (–301 to 364) | 0.896 |
| Medication use * | 108 (–281 to 632) / –17 (–299 to 292) | 0.136 | 82 (–256 to 517) / –84 (–371 to 226) | <0.00 |

\*Yes/no are shown.

**Table 4.**
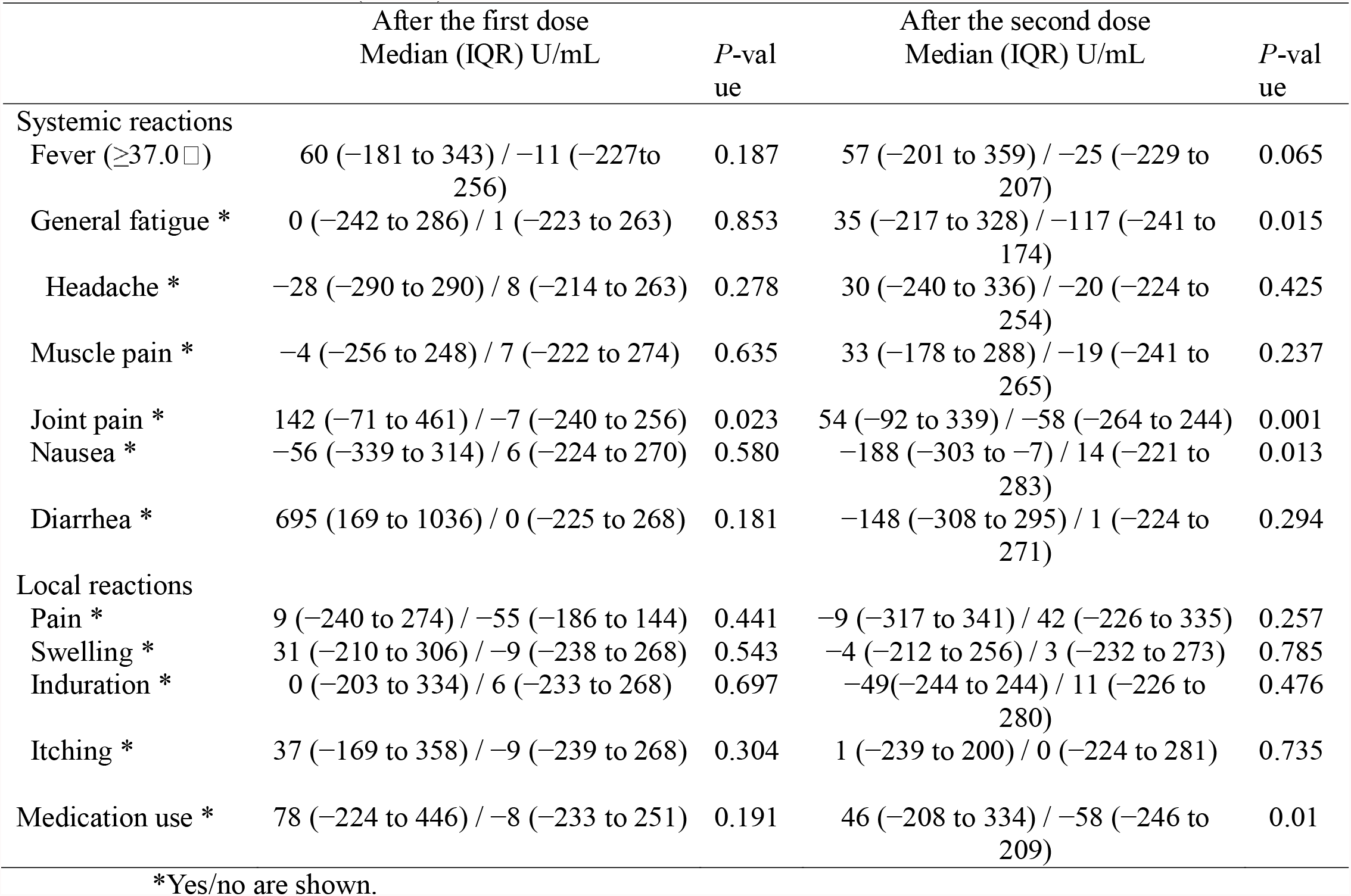
Relationship between adverse effects and age-adjusted antibody titers at 6 months after vaccination (n=365)

**Figure 2.**
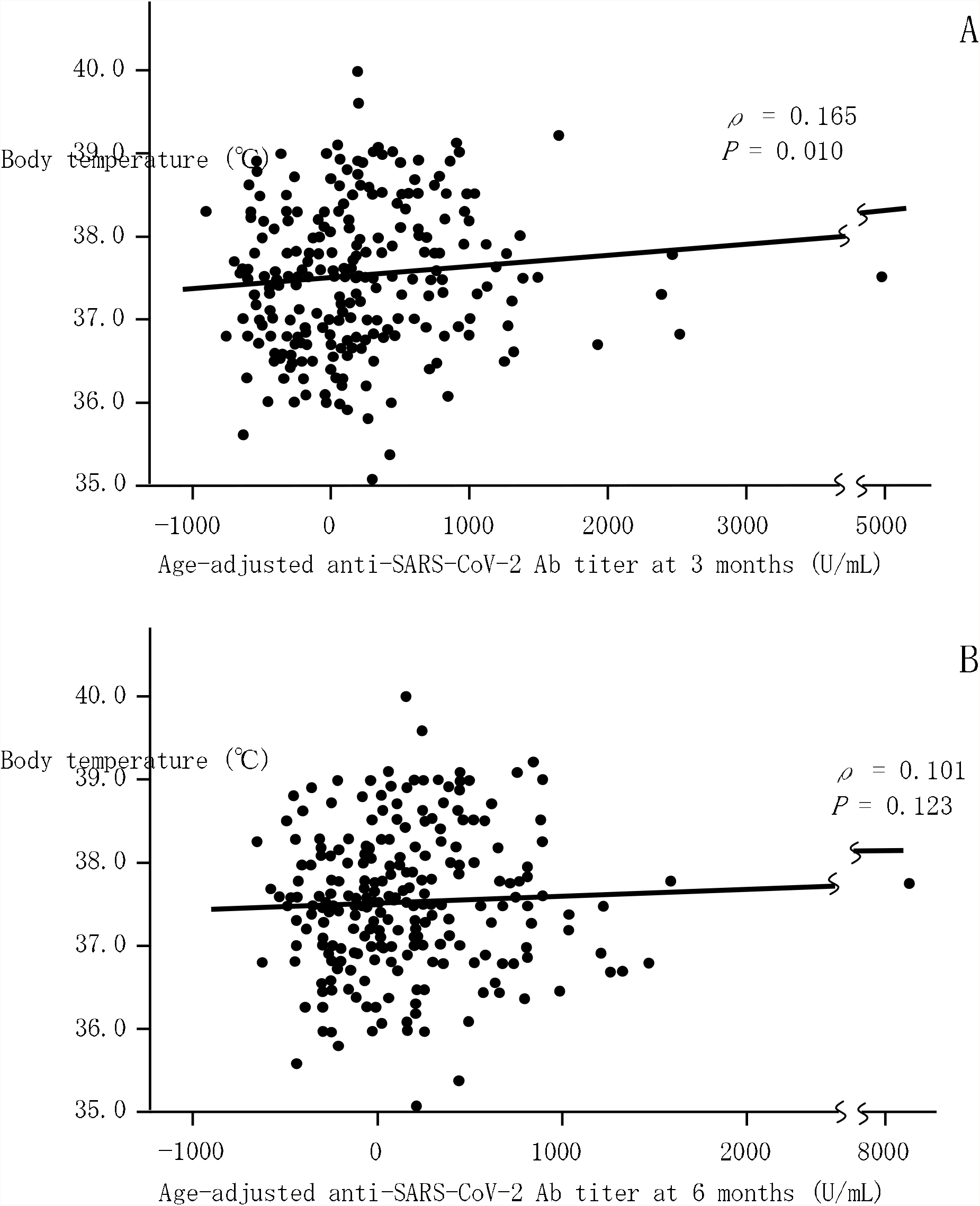
Relationships between body temperature after the second vaccine dose and age-adjusted anti-SARS-CoV-2 Ab titers at 3 months (A) and 6 months (B) after vaccination. Significant positive correlations were observed at 3 months after vaccination, but not at 6 months.

The differences in age-adjusted Ab titers at 3 months after vaccination between individuals with and without fever after the first and second doses were 181 U/mL and 179 U/mL, respectively. These values were 23.7% and 23.4% of the median Ab titer of 746 U/mL at 3 months after vaccination. In addition, differences in the age-adjusted Ab titers between individuals with and without anti-inflammatory agent use after the second vaccine dose were 166 U/mL and 104 U/mL at 3 and 6 months after vaccination, respectively. These values were 21.7% and 19.3% of the median Ab titers of 746 U/mL and 539 U/mL at 3 and 6 months after vaccination, respectively.

## DISCUSSION

To our knowledge, this is the first study to report the relationship between adverse effects and Ab titers against the SARS-CoV-2 spike antigen from 3 to 6 months after vaccination with the BNT162b2 vaccine in Japan. Three important findings were obtained. First, while injection-site symptoms occurred with almost equal frequency for the first and second vaccine doses, systemic reactions were significantly more common after the second dose (**Table 1**). General fatigue and fever were the first and second most common systemic adverse effects. Second, multivariate analysis revealed significant correlations of fever with female sex for the second dose, younger age for both the first and second doses, and dyslipidemia for the first dose (**Table 2**). No other clinical or lifestyle characteristics were correlated with fever. Third, regarding the relationship between systemic adverse effects and Ab titers (**Tables 3 and 4**), participants with general fatigue after the second dose had significantly higher age-adjusted Ab titers at both 3 months and 6 months than those without general fatigue. Compared with participants without fever, those with fever after the first or second dose had significantly higher age-adjusted Ab titers at 3 months, but not at 6 months. Participants with joint pain after the first or second dose had significantly higher age-adjusted Ab titers at both 3 months and 6 months than those without joint pain. Participants who used anti-inflammatory agents after the second dose showed a significantly higher Ab titer.

Not only high Ab responses against SARS-CoV-2 spike protein, but also systemic adverse effects induced by the BNT162b2 vaccine were reported to be more common among participants with preexisting immunity due to COVID-19 infection [10], suggesting that some systemic adverse effects are induced as a hypersensitive immune reaction to the SARS-CoV-2 spike protein itself produced by the BNT162b2 vaccine. Previous publications reported that adverse effects induced by the BNT162b2 vaccine had no or only a low correlation with Ab titers detected immediately after vaccination [5-8]. However, to evaluate the long-term effects of vaccination, the research should consider the Ab titers several months after vaccination, not just the peak level of Ab titers. Accordingly, we determined the relationship between adverse effects and Ab titers from 3 to 6 months after the second dose of the BNT162b2 vaccine. Few studies have addressed this issue, but we found one report from Estonia showing that the Ab response was negatively correlated with age and positively correlated with the total score for vaccination adverse effects [9].

For the second findings, we found significant correlations of female sex, younger age, and dyslipidemia with fever (**Table 2**). Some studies have reported that women and younger people more commonly show adverse effects of the BNT162b2 vaccine [6], and our results support those findings. However, why dyslipidemia would be a risk factor for fever is not known. Atherogenic dyslipidemia may have effects on the activity of collagen diseases, such as systemic lupus erythematosus and rheumatoid arthritis [14, 15]. Diabetes mellitus, a known immune disorder, was also a near significant risk factor for adverse effects after the first dose. These metabolic diseases complicate immune disorders, and these immune disorders may contribute to systemic reactions after the first vaccine dose.

Regarding the third finding, the occurrence of systemic adverse effects, including general fatigue, fever, and joint pain, and the use of an anti-inflammatory agent for adverse effects were significantly associated with a high Ab titer. In particular, body temperature recorded as an adverse effect was significantly and positively correlated with the Ab titer at 3 months after vaccination, suggesting that the severity of systemic adverse effects may promote a stronger ability to maintain high Ab titers against the spike protein of this virus. Joint pain contributed to significantly higher Ab titers after both the first and second doses at both 3 and 6 months after vaccination. Joint pain was one of the most common reactions. However, we could not find previous publications that explained the relationship between Ab titer and joint pain, and we also are unable to speculate why Ab titers were more strongly reflected by joint pain than by fever or general fatigue.

Some limitations and possible sources of bias in this study include the following. First, the participants were limited in number and were all healthcare workers vaccinated at a single national hospital in Tochigi Prefecture. Therefore, the results obtained in this study might not be widely generalizable or even generalizable within Japan. Second, we may need additional analysis and discussion to determine whether age is an important factor independently associated with adverse effects. One possibility is that fever may be an independent factor associated with higher Ab titers, but not age. In this case, because most of the participants with fever were younger, younger age may not be independently associated with a higher Ab titer. The opposite is also a possibility. In addition, both fever and age may be important factors. Therefore, we cannot determine whether fever or young age is a more important factor.

In conclusion, participants with systemic adverse effects had about 20% higher Ab titers at 3 and 6 months after the second dose of the BNT162b2 vaccine. Although the systemic adverse effects induced by the BNT162b2 vaccine are sometimes unpleasant, higher Ab titers should reduce the risk of a more severe COVID-19 infection. We hope that our results will encourage vaccine uptake.

## Data Availability

All data produced in the present work are contained in the manuscript

## Author Contributions

Conceptualisation, M.S., K.S., Y.N. (Yushi Nomura) and Y.N. (Yosikazu Nakamura); methodology and software, Y.N. (Yushi Nomura), K.S., M.S. and Y.N. (Yosikazu Nakamura); validation, K.S., M.S. and Y.N. (Yosikazu Nakamura); formal analysis and investigation, K.S., Y.N. (Yushi Nomura), M.S. and Y.N. (Yosikazu Nakamura); resources, T.T., K.S., N.M., Y.N. (Yushi Nomura), M.S., O.K., and R.K.; data curation, Y.N. (Yushi Nomura), K.S. and M.S.; writing—original draft preparation, M.S., K.S., Y.N. (Yushi Nomura) and Y.N. (Yosikazu Nakamura); writing—review and editing, T.T., N.M., S.N., K.H., O.K., and R.K.; visualisation, K.S., Y.N. (Yushi Nomura) and M.S.; supervision, M.S., K.S., T.T., S.N. and K.H.; project administration, K.S. and T.T.; funding acquisition, T.T. and K.S. All authors have read and agreed to the published version of the manuscript.

## Institutional Review Board Statement

The study was conducted according to the guidelines of the Declaration of Helsinki and approved by the Ethics Committee of National Hospital Organization Utsunomiya National Hospital (No. 03-01, 19 April 2021).

## Informed Consent Statement

Informed consent was obtained from all subjects involved in the study.

## Acknowledgments

The authors thank all staff at National Hospital Organization Utsunomiya National Hospital for providing support during sample collection. We would like to thank Miwa Okada, Minako Yamagishi, Junko Shibayama, Yuko Tajima, Mami Ochiai, Midori Takahashi, Hiroko Ueno, Natsuka Suzuki and Yoshino Iwaya for providing support during data analysis. Conflicts of Interest: The authors declare no conflict of interest.

## Abbreviations

COVID-19: coronavirus disease 2019
Ab: antibody
SARS-CoV-2: severe acute respiratory syndrome coronavirus 2
ECLIA: electrochemiluminescence immunoassay
IQR: interquartile range
OR: odds ratio
CI: confidence interval

